# A novel modelling framework to simulate the effects of HIV stigma on HIV transmission dynamics

**DOI:** 10.1101/2024.10.01.24314728

**Authors:** Donal Bisanzio, Sarah T. Roberts, Rachel D. Stelmach, Kendall N. McClellan, Georgiy Bobashev, Joella Adams, Katherine Karriker-Jaffe, Stacy M. Endres-Dighe, Khalida Saalim, Natalie Blackburn, Laura Nyblade

## Abstract

**Introduction:** HIV remains a global public health challenge, with social determinants such as stigma influencing transmission dynamics, access to testing, and treatment. HIV stigma shapes both individual behaviour and community responses to HIV. However, modelling approaches have rarely represented the complex role of stigma in HIV epidemics. Our study introduces an innovative modelling framework designed to capture the interplay between stigma and HIV transmission dynamics.

**Methods:** We modelled effects of anticipated, internalised, and experienced HIV stigma on HIV testing, antiretroviral treatment initiation, and treatment adherence. We built an individual-based model representing the HIV epidemic (HIV-IBM) in a USA-like population of 3 million individuals that accounts for community demography, behaviour, and healthcare access. Stigma parameters were based on a scoping review focused on the prevalence and effects of stigma in people living with and without HIV. HIV-IBM was used to assess effects of interventions targeting different types of stigma. We tested reductions of stigma by 50% and 100% across the simulated population and performed a sensitivity analysis to identify effects of each type of stigma on the simulated HIV epidemic.

**Results:** Without reduced stigma, the HIV-IBM had an annual incidence rate of 12.6 (95% credible interval [CI]: 11.4-13.5) new cases per 100,000 people. Reducing the overall level of stigma in the population by 50% resulted in an annual incidence rate of 9.6 (95% CI: 8.6-10.3) per 100,000, and a 100% reduction in stigma resulted in an annual incidence rate of just 6.8 (95% CI: 6.1-7.3) per 100,000. In addition to reducing HIV incidence, reducing stigma resulted in a substantial increase of viral suppression among people living with HIV (50% stigma reduction: +10.5%; 100% stigma reduction: +16.4%). Sensitivity analysis showed that outcomes resulting from interventions targeting each type of stigma were highly heterogeneous.

**Conclusion:** Simulation results suggest that reducing HIV stigma could have a large effect on HIV incidence and viral suppression. Our model framework provides a dynamic approach to understanding the role of stigma in HIV outcomes that facilitates exploration of stigma reduction strategies and offers insights to inform evidence-based policies and interventions for reducing stigma and curtailing HIV.

## INTRODUCTION

HIV remains a global public health challenge, with over 39 million people living with HIV (PLWH) and about 1.3 million new cases globally in 2022, including 31,800 new cases in the United States (US).(1) Both the UNAIDS Global AIDS Strategy and U.S. *Ending the HIV Epidemic* Initiatives have set targets of 95% of PLWH aware of their status, 95% linkage to care among those diagnosed with HIV, and 95% viral load suppression among those linked to care by 2025.(2,3) Despite some countries’ success, many more are unlikely to meet these targets. In the US in 2022, 87% of PLWH were aware of their status, and among those, 83% have been linked to care and 65% were virally suppressed.(4)

HIV stigma shapes both individual behaviour and community responses to HIV, as well as transmission dynamics and utilisation of testing and treatment. As described by Turan and colleagues (5), forms of HIV stigma can include enacted or experienced HIV stigma, which occurs when people experience actual discriminatory actions due to their HIV status; perceived HIV stigma, which describes the negative attitudes a person believes other people hold about PLWH; anticipated HIV stigma, which occurs when a person expects that other people will treat them badly due to their HIV status; and internalised HIV stigma, which occurs when PLWH accepts that negative stereotypes about PLWH are true and applicable to themself. These types of HIV stigma have been demonstrated to influence uptake and adherence to highly effective interventions such as HIV testing, antiretroviral therapy (ART), and pre-exposure prophylaxis (PrEP), as well as HIV-related health outcomes such as viral load suppression, although results vary across study designs, measures, and populations.(6–13)

Mathematical models can synthesize key evidence for understanding the HIV epidemic; informing policy decisions; predicting long-term health outcomes; and characterizing the interaction of biological, social, and behavioural factors to reduce or exacerbate those outcomes. Despite the known influence of stigma on HIV transmission and progression, HIV models have rarely included stigma.(14–19) Modellers choose to exclude stigma due to perceptions that stigma is too complex, poorly understood, or difficult to measure to implement in a modelling process.(14,17,18) Not accounting for stigma when estimating the effectiveness of HIV-related interventions omits important public health considerations and prevents policymakers from fully evaluating the potential impact of stigma-reduction interventions. These interventions may have greater benefit than expected due to the cumulative effect of stigma on multiple steps of the prevention and treatment cascade. In this mathematical modelling study, we introduce an innovative framework designed to estimate the effects of anticipated, internalised, and experienced HIV stigma on HIV testing, treatment, and transmission dynamics.

## METHODS

We estimated the impact of interventions targeting stigma using an individual based model (HIV-IBM) representing a synthetic population of a hypothetical city of 3 million people in the US. We provide an overview of the HIV-IBM here, with a more detailed description of the model provided in the Appendix. The HIV-IBM simulates HIV transmission through two main HIV transmission routes: syringe sharing among people who inject drugs (PWID) and sexual intercourse among serodiscordant partners. Thus, the HIV-IBM includes two contact networks, with one representing the sexual network and another capturing needle sharing among PWID. The HIV-IBM’s sexual network simulates sexual contacts among individuals above 14 years of age in the USA. This network accounts for sexual contact of men who have sex with women (MSW), women who have sex with men (WSM), men who have sex with men (MSM), women who have sex with women (WSW), men who have sex with both men and women (MSMSW), and women who have sex with both women and men (WSWSM). This approach assumes that transgender individuals identify as either a man or a woman and notably omits behaviours of individuals of any other gender identity (e.g. nonbinary people). The HIV-IBM’s needle-sharing network uses a contact network to represent the action of sharing syringes among people who inject drugs (PWID), with a proportion of the network assigned as HIV positive at the start of the simulation (see details below and in Appendix 1). The network of needle sharing was created to represent needle sharing behaviour among PWID in the US.(20) The HIV-IBM also simulates HIV testing and treatment in the synthetic population. Proportions of people who get tested for HIV, PLWH who start treatment, and PLWH who adhere to treatment regimens were based on data collected from published studies (Appendix 1, Table A5). The sexual network resulted from individuals within the model interacting as they moved across the simulated city. Their movement was simulated using a gravity model that has previously been used to model Ebola, COVID-19, and mpox.(21–23)

The HIV-IBM aimed to assess the effects of reducing HIV stigma on HIV transmission. Thus, the HIV-IBM simulates the impact of stigma at several points, as stigma can prevent people from getting tested, delay the start of treatment, and reduce the probability of treatment adherence. We modelled three different manifestations of stigma: anticipated stigma, internalised stigma, and experienced stigma (Table 1).(24)

**Table 1:** Types of stigma modelled.

| Stigma type | Affected behaviour | Source used in model |
| --- | --- | --- |
| Anticipated stigma | Getting tested | (25) |
|  | Adhering to ART | (26) |
| Internalised stigma | Starting ART | (27) |
|  | Adhering to ART | (26) |
| Enacted Stigma | Adhering to ART | (26) |

Studies investigating the effects of stigma typically use continuous scales to represent level of stigma affecting the target population rather than dichotomous variables (“affected by stigma” or “not affected by stigma”).(28) Thus, the HIV-IBM included stigma using scale scores from different sources on the effects of stigma on the HIV care continuum. Given the absence of a single standardized scale to quantify the level of stigma types, we adopted scales reported in the studies from which we gathered information about the associations of stigma with testing, starting treatment, and adhering to treatment (Appendix 1, Table A3).

The parameters used to build the HIV-IBM starting population and its characteristics are reported in the Appendix. The model simulation runs did not begin with a HIV-naïve population, but rather initiated with a population representing the US population with current levels of HIV prevalence. We disaggregated the number of cases in the starting population across sexual orientation, race, and ethnicity (Appendix 1, Table A5) (29–31). The starting populations in the HIV-IBM also included a fraction of non-tested individuals, PLWH who know their status but have not started HIV treatment, and PLWH who are not adherent to HIV treatment (Appendix 1, Table A5). The IBM-HIV simulated HIV transmission over a three-year time window. Weekly probabilities of HIV testing, initiating ART after a positive HIV test, and ART adherence were estimated at the individual level.

The baseline model was calibrated to reflect HIV incidence reported for 2022, approximately 11.3 infected every 100,000 people (32). Results are reported using the median and the 95% credible interval (CI, as 2.5 and 97.5 quantile) based on 10,000 simulations per scenario. We evaluate the effect of stigma on HIV transmission by simulating scenarios in which we changed the level (i.e., score) of each manifestation of stigma throughout the population. We compare the baseline model with two scenarios:

- Anticipated, internalised, and experienced stigma reduced by 50%
- Eliminating anticipated, internalised, and experienced stigma (i.e., simulated effects of 100% stigma reduction)

We performed a sensitivity analysis to quantify the effect of anticipated, internalised, and experienced stigma on the HIV care continuum and HIV transmissions. A full description of the sensitivity analysis is provided in the Appendix.

## RESULTS

The baseline model had an average annual HIV incidence of 12.6 (95% credible interval [CI]: 11.4-13.5) new cases per 100,000 people, with 379 (95% CI: 242-405) incident HIV cases per year and 1,141 (95% CI: 931-1,350) incident HIV cases over the three-year time window (Figure 1, Figure 2, and Table 2). The modelled scenario reducing levels of anticipated, internalised, and experienced HIV stigma by 50% resulted in annual incidence of 9.6 (95% CI: 8.6-10.3) new cases per 100,000 people, an average of 288 new cases per year (95% CI: 258-329), and an average of 872 (95% CI: 594-596) new cases at the end of the three years (Figure 1, Figure 2, and Table 2). Eliminating anticipated, internalised, and experienced stigma resulted in an annual incidence of 6.8 (95% CI: 6.1-7.3) new cases per 100,000 people, an average of 204 (95% CI:: 183-219) new cases per year, and an average of 612 (95% CI: 521-671) new cases at the end of the three years equal to (Figure 1, Figure 2, and Table 2).

**Figure 1:**
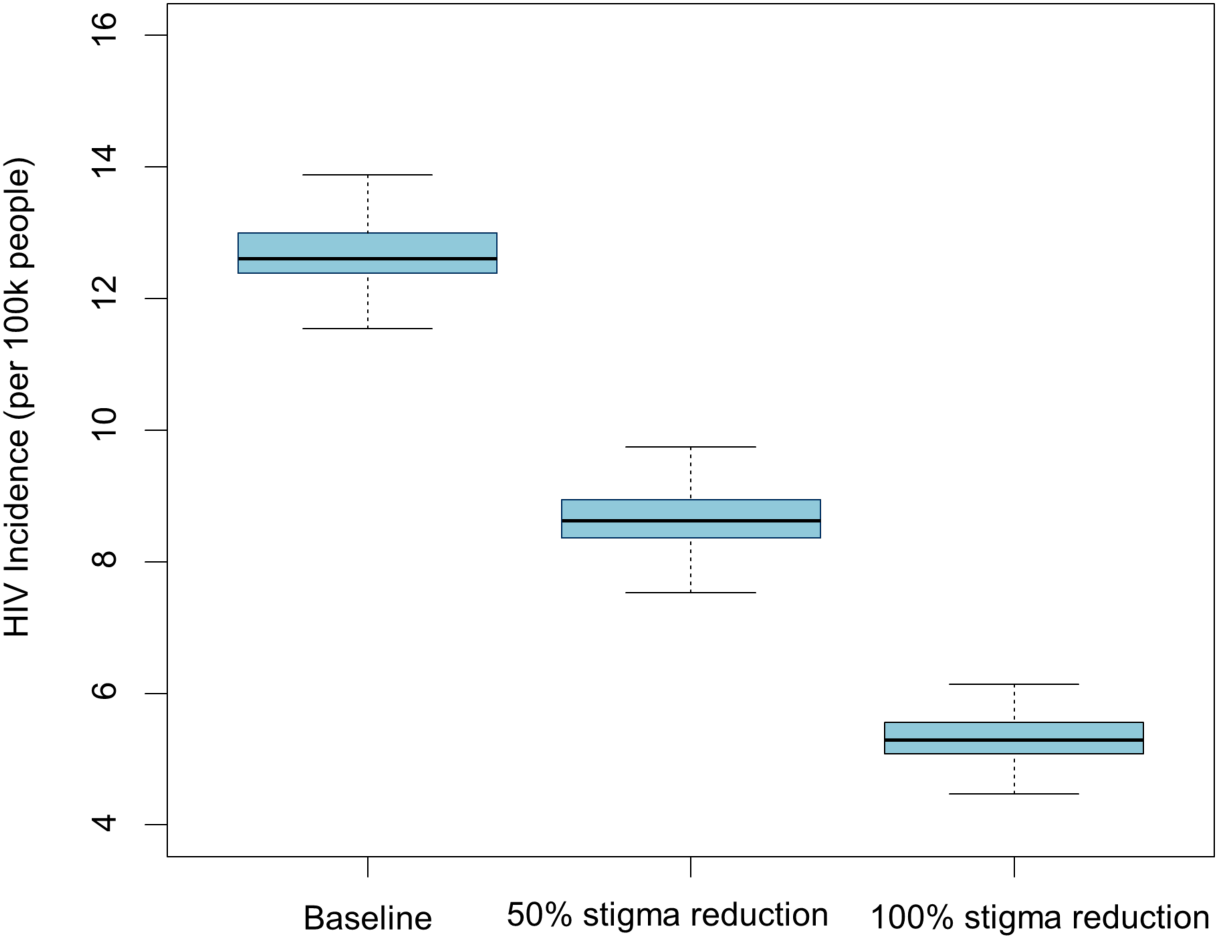
Effect of stigma reduction on HIV incidence.

**Figure 2:**
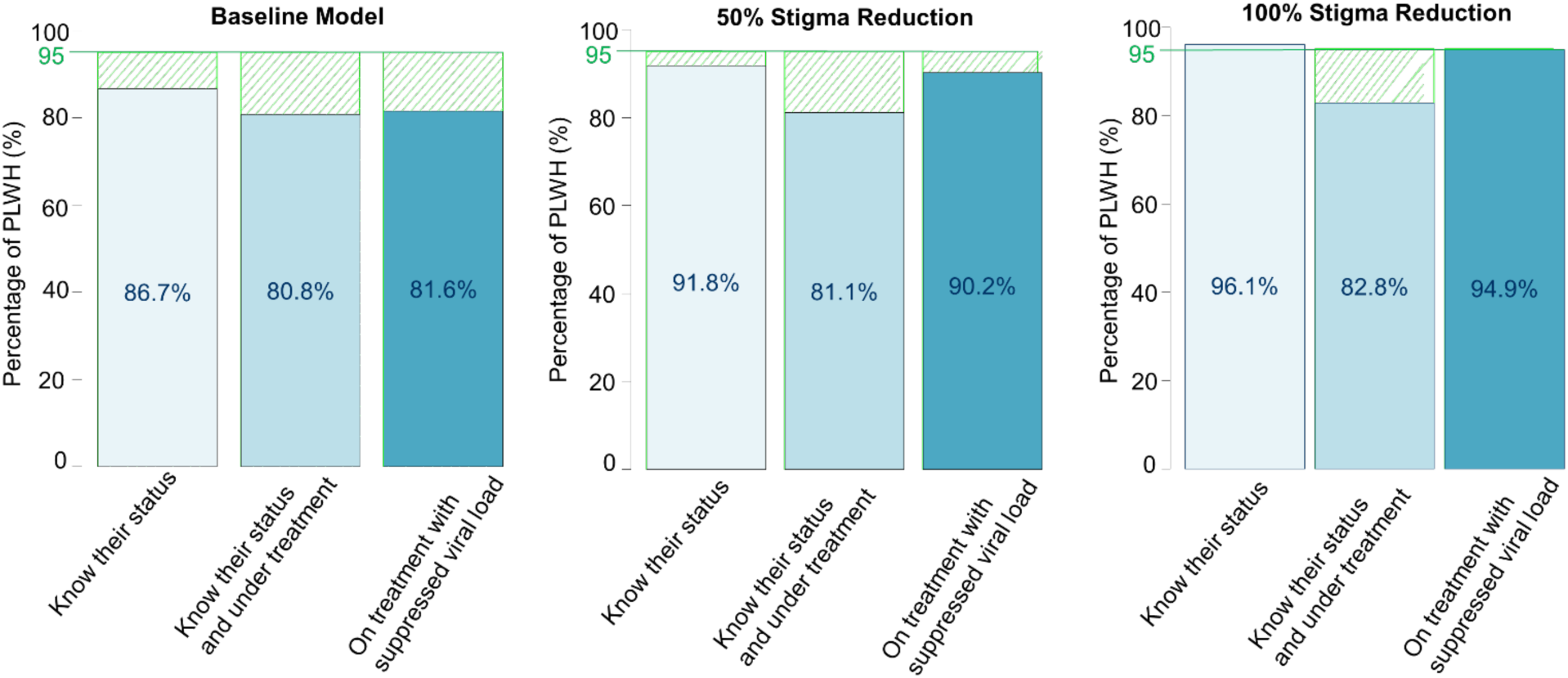
Effect of stigma reduction on HIV testing, treatment, and viral load suppression: Comparison to UNAIDS 95-95-95 targets.

**Table 2:** Results of the three scenarios simulated with the HIV-IBM.

| Scenario | Baseline |  | HIV stigma reduced by 50% |  | HIV stigma reduced by 100% |  |
| --- | --- | --- | --- | --- | --- | --- |
|  | Estimate | 95% CI* | Estimate | 95% CI | Estimate | 95% CI |
| Mean annual incidence per 100,000 people | 12.6 | 11.4 -13.5 | 9.6 | 8.6-10.3 | 6.8 | 6.1-7.3 |
| Mean new cases per year | 379 | 342-405 | 288 | 258-329 | 204 | 183-219 |
| Total number of new cases | 1,141 | 931-1,350 | 872 | 594-956 | 612 | 521-671 |
| Percentage of PLWH knowing their status | 86.7% | 82.1-90.1% | 91.8% | 87.2-94.1% | 96.1% | 91.3-98.2% |
| Percentage of PLWH who know their status who are on treatment | 80.8% | 75.5-86.3% | 81.1% | 73.2-85.3% | 90.2% | 87.7-93.3% |
| Percentage of PLWH on treatment with suppressed viral load | 81.6% | 76.1-88.6% | 90.2% | 87.7-93.3% | 94.9% | 90.3-97.4% |

\*CI: credible interval

Stigma reduction had significant impacts throughout the HIV care continuum. Reducing stigma increased the proportion of PLWH who know their status and the proportion of people on treatment who had achieved viral load suppression (Table 2 and Figure 2). The fraction of PLWH who know their status increased from 86.7% (95% CI: 82.1%-90.1%) to 91.8% (95% CI: 87.2%-94.1%) when stigma was reduced by 50%, and to 96.1% (91.3%-98.2%) when stigma was eliminated. Eliminating stigma had a slightly positive effect on HIV treatment initiation for PLWH with an HIV diagnosis: a 2.5% increase (baseline: 80.8% [95% CI: 75.5%-86.3%] of PLWH with an HIV diagnosis who initiated treatment, and 100% reduction: 82.8% [95%: 78.5%-92.8%] who initiated treatment) (Table 2, and Figure 2). Viral suppression among PLWH under treatment drastically increased comparing the baseline model and those with reduced or eliminated stigma. Compared to viral suppression resulted obtained from the baseline scenario (81.6% [95% CI: 76.1%-88.6%]), the HIV-IBM with 50% stigma reduction resulted in a 10.5% increase in viral suppression (90.2% [95% CI: 87.7%-93.3%]) among treated PLWH. Eliminating HIV stigma resulted in a 16.2% increase compared to baseline (94.9% [95% CI: 90.3%-97.4%]). The sensitivity analysis showed that reducing anticipated stigma affecting HIV testing provided the highest mean reduction of HIV incidence compared to other stigma effects (Figure 3). However, sensitivity analyses also showed that reducing experienced stigma affecting ART adherence had little effect on HIV incidence (Figure 3).

**Figure 3:**
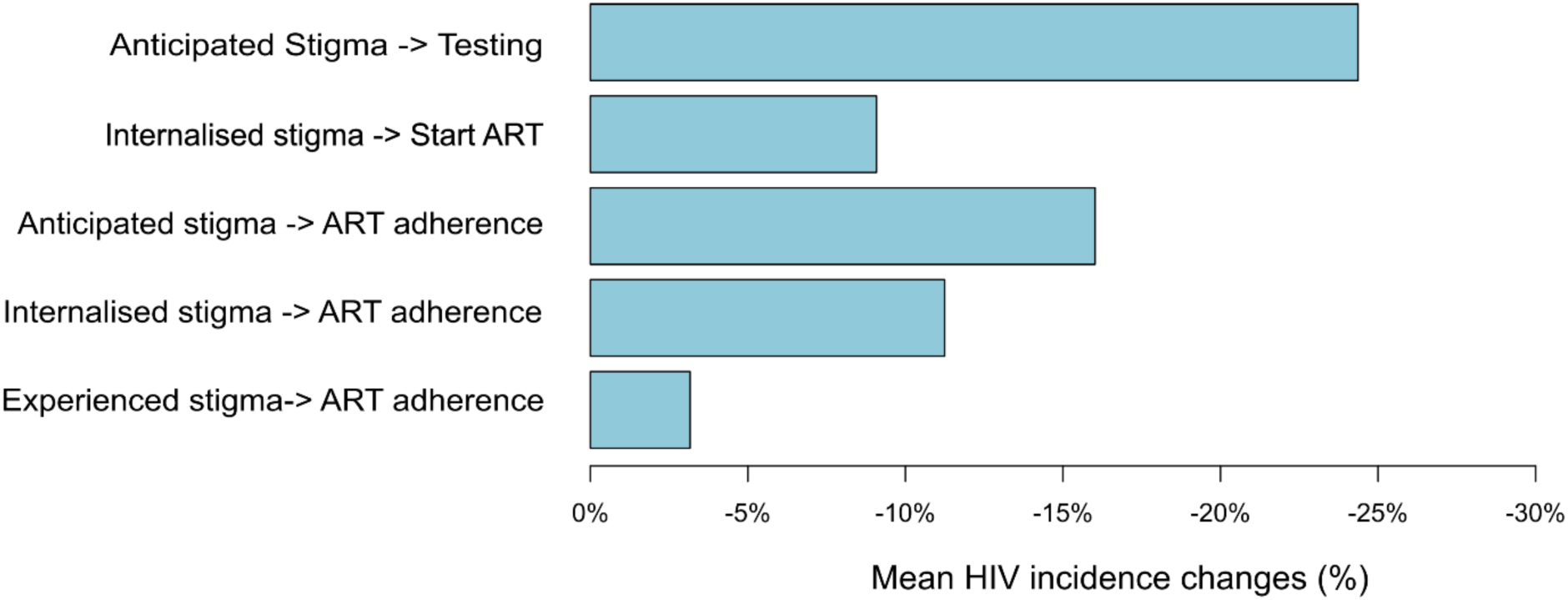
Effects of 100% reduction in individual types of stigma.

## DISCUSSION

This paper introduces a framework for incorporating HIV stigma into mathematical models of the HIV epidemic. Findings suggest that eliminating HIV stigma could have a large impact on HIV incidence and make substantial progress towards the 95-95-95 UNAIDS targets, leading to achievement of the 1^st^ 95% target and near-achievement of the 3^rd^ target. Our results also suggest that interventions to reduce anticipated stigma’s effect on HIV testing could have the greatest single impact on HIV incidence of the included stigma-reduction interventions. While our point estimates and their simulation bound intervals are limited to the model and underlying synthetic population in a hypothetical U.S. city, the results show an expected effect of stigma reduction that provides an initial estimate of the effects of potential interventions. Application of this model’s novel, dynamic approach in real-world settings, and to specific subpopulations who experience higher levels of stigma, could strengthen the case for investment in stigma reduction efforts; facilitate the exploration of specific, tailored stigma reduction strategies; and offer insights to inform evidence-based policies and programming to reduce HIV stigma and curtail the epidemic.

We are aware of three other mathematical models that have investigated the effects of stigma on the HIV epidemic. Stover et al. incorporated internalised HIV stigma into a deterministic compartmental HIV epidemic simulation model for each of 77 countries with a high burden of HIV.(19) Based on a hypothetical scenario assuming that the 95-95-95 goals can be achieved only in the absence of internalised stigma, they found that failure to reduce internalised stigma would substantially increase both HIV incidence and HIV-related mortality by inhibiting rates of HIV testing, treatment, and viral load suppression. Because their model does not include anticipated or enacted stigma, it likely underestimates the potential impact of stigma elimination. Prudden et al. used a probabilistic mathematical model to estimate the impact of stigma on vertical transmission of HIV in South Africa.(18) Under the scenario of universal access to ART for all pregnant women, the model predicted that eliminating stigma would avert 51% of all infant HIV acquisitions, with the largest impact on the treatment adherence stages of the cascade.(18) Their model does not disaggregate various forms of stigma to guide intervention choices.

Further, by nature of its focus on pregnant women, the results are not easily translated to other populations with different levels of engagement with the healthcare system and different motivations for treatment.(33) Our model had results similar in magnitude to these two preceding models, but it expands upon their work by separately modelling different forms of stigma at different stages of the prevention and care cascade to allow more detailed and comprehensive estimates of the potential results of stigma reduction efforts.

In contrast to Prudden et al. and Stover et al., Levy et al. estimated the effects of HIV-related stigma on epidemic dynamics in Kenya by integrating a mechanistic model of HIV stigma—based on prevalence estimates of enacted and perceived stigma from the Demographic and Health Surveys and assumptions on stigma dynamics—into a compartmental of HIV transmission dynamics.(17) Levy et al. found that eliminating enacted stigma between 2004 and 2017 would have reduced HIV incidence from about 5% to 3%, while eliminating internalised stigma would have had little impact on incidence. Additionally, they found that stigma elimination would have only a small impact on annual HIV incidence and HIV-related mortality by 2030.(17) Key limitations to the model of Levy et al. are its reliance on fitted formulas rather than epidemiologic data to estimate the prevalence of internalised stigma and the impact of stigma on HIV outcomes, which may account for the differences in this model’s findings compared to our own and to those of Stover et al. and Prudden et al.(17–19)

The quantity and nature of the available data on HIV stigma posed significant challenges in parameterizing the model. While data on the prevalence or severity of HIV stigma are plentiful, few studies have tested the impact of stigma on HIV-related health outcomes, and those studies have shown inconsistent results. A possible explanation for these inconsistencies is the lack of standard definitions and measures for different forms of HIV stigma. We chose to model anticipated, enacted, and internalised stigma which are among the most commonly studied forms of stigma, but other researchers have also reported on *community* stigma, *public* stigma, *felt* stigma, *self* stigma, *structural* stigma, *professional* stigma, and other forms. Further, the definitions and items used to measure these various forms of stigma often overlap. For example, disclosure concerns have been classified as representing either internalised or anticipated stigma, depending on the scale (34–36). When consensus exists on the forms and definitions of stigma, multiple scales are still used to quantify stigma, which makes it difficult to compare research results and use them to parametrize a model. There was also insufficient data to model interactions between different types of stigma, such as the potential for internalized stigma to mediate effect of experienced stigma on medication adherence.(37) Finally, studies use various techniques to model stigma as an exposure, such as continues scores, z-scores, threshold levels, or percentiles for the study population.. This variation makes results difficult to aggregate or compare across studies. These challenges point to the need for greater consensus in stigma definitions, a more consistent approach to analysing stigma across studies, and more well-designed studies to evaluate the impact of HIV-related stigma on health outcomes, as well as dedicated resources and leadership to accomplish these objectives.

For better and worse, the HIV-IBM model reflects the current state of HIV stigma measurement at the level of the full population. The model’s use of continuous scales for stigma instead of a dichotomous “stigma” or “no stigma” model allows it to describe heterogenous experiences of stigma across the population. It also provides insight into the effects of different manifestations of stigma on different aspects of HIV prevention and treatment, which allows for a more nuanced understanding of how stigma affects HIV transmission. The flexible nature of the model also makes it easy to adjust for future work that provides further detail on specific populations, conditions, and interventions. The current model represents what the authors feel is a reasonable balance between fidelity (complex but true-to-life) and tractability (simple but feasible to program), but as the availability of data improves, the HIV-IBM should be also revisited and improved. The current iteration of the model does not capture intersectional stigma, nor does it describe differences in stigma affecting people of different marginalized identities, e.g. by race, ethnicity, gender, sexual orientation, etc. Importantly, while the current model includes people with multiple patterns of sexual behaviour, it does not include people with gender identities other than “man” or “woman”. Future work building upon and expanding the HIV-IBM should consider these additional variables as means for improving the fidelity and specificity of the model, and thereby increasing the model’s usefulness for understanding the effects of HIV stigma on the populations most affected by HIV.

Both UNAIDS and the U.S. National HIV/AIDS Strategy frame stigma as a societal barrier to achieving the 95-95-95 targets, and have explicit goals to reduce stigma experienced by PLWH and people disproportionately affected by HIV. (2,3) Our results suggest that eliminating or reducing stigma will, in fact, help to attain the 95-95-95 targets. Multiple interventions have been shown to effectively reduce HIV stigma among people living with HIV, healthcare workers, and the community (38–45), including education, contact strategies, counselling, and structural changes to laws and policies.(39) While not explicitly included in the current model, evidence suggests that stigma reduction may also improve other health outcomes, such as depression, anxiety, social support, access to health and social services, and overall quality of life.(46) Future iterations of this model could incorporate scenarios mimicking specific evidence-based stigma reduction interventions at different levels of the health system and of society and incorporate additional health outcomes to understand the broader impact of stigma reduction on the health and well-being of PLWH. Although our model suggests that reducing anticipated stigma would have the greatest impact on HIV testing and incidence, there are fewer evidence-based interventions to reduce this form of stigma. Multiple studies show community members’ negative attitudes towards people living with HIV have declined over time, but this is not always accompanied by declines in anticipated stigma, suggesting that to effectively reduce anticipated stigma, additional steps must be taken to convince individuals that others in the society have also changed their views.(47,48)

## CONCLUSIONS

Our model framework provides a dynamic approach to understanding the role and estimating the effect of stigma in HIV transmission. Findings suggest that reducing HIV stigma could have a large impact on HIV incidence and strengthen the case for investment in actions to reduce stigma and discrimination. This model’s novel approach could facilitate the exploration of stigma reduction strategies and offer insights to inform evidence-based policies and interventions for reducing stigma and curtailing HIV.

## AUTHORS’ CONTRIBUTIONS

DB, STR, GB, and LN conceptualized the research. KNM, RDS, and DB curated data inputs for the model. DB developed the model and conducted the formal analysis. DB, STR, RDS, KNM, LN, JA, KKJ, SMED, NB, and GB participated in model structuring and parameterization. LN and GB secured funding for this project and provided supervision. STR, LN, and RDS provided project administration support. DB, STR, and RDS wrote the first draft of the manuscript and GB, JA, LN, KNM, SMED, KS, NB, and KKJ provided substantial revisions. All authors read and approved the final manuscript.

## ACKNOWLEDGEMENTS

The authors thank members of the RTI Stigma Community of Practice for their contributions to the model design and are grateful to the participants who contributed data to the many studies used to parameterize the model.

## DATA AVAILABILITY STATEMENT

Data sharing not applicable – no new data generated

## SUPPORTING INFORMATION

## Appendix 1: Technical appendix

Microsoft Word document. Contains details of parameter values and their sources as well as a more detailed description of the modelling methods.

## LIST OF ABBREVIATIONS

ART: Antiretroviral therapy
CI: Credible interval
HIV: Human immunodeficiency virus
HIV-IBM: Individual-based model representing the HIV epidemic
MSM: Men who have sex with men
MSMSW: Men who have sex with both men and women
MSW: Men who have sex with women
WSM: Women who have sex with men
WSW: Women who have sex with women
WSWSM: Women who have sex with both women and men
PrEP: Pre-exposure prophylaxis
PLWH: People living with HIV
PWID: People who inject drugs

## REFERENCES

1. Centers for Disease Control and Prevention. HV Surveillance Supplemental Report: Estimated HIV Incidence and Prevalence in the United States, 2018–2022. 2024 May 21; Available from: https://stacks.cdc.gov/view/cdc/156513

2. UNAIDS. Global AIDS Strategy 2021-2026. End Inequalities. End AIDS. [Internet]. Geneva, Switzerland; 2021 [cited 2024 Sep 16]. Available from: https://www.unaids.org/en/resources/documents/2021/2021-2026-global-AIDS-strategy

3. The White House. National HIV/AIDS Strategy for the United States, 2022-2025. Washington, DC; 2021.

4. Centers for Disease Control and Prevention. HV Surveillance Supplemental Report: Monitoring Selected National HIV Prevention and Care Objectives by Using HIV Surveillance Data United States and 6 Territories and Freely Associated States, 2022. 2024 May 21; Available from: https://stacks.cdc.gov/view/cdc/156511

5. Turan B, Hatcher AM, Weiser SD, Johnson MO, Rice WS, Turan JM. Framing mechanisms linking HIV-related stigma, adherence to treatment, and health outcomes. Am J Public Health. 2017 Jun;107(6):863–9.

6. Rueda S, Mitra S, Chen S, Gogolishvili D, Globerman J, Chambers L, et al. Examining the associations between HIV-related stigma and health outcomes in people living with HIV/AIDS: a series of meta-analyses. BMJ Open. 2016 Jul 1;6(7):e011453.

7. Katz IT, Ryu AE, Onuegbu AG, Psaros C, Weiser SD, Bangsberg DR, et al. Impact of HIV-related stigma on treatment adherence: systematic review and meta-synthesis. J Int AIDS Soc. 2013 Nov 13;16(3 Suppl 2):18640.

8. Christopoulos KA, Neilands TB, Dilworth S, Lisha N, Sauceda J, Mugavero MJ, et al. Internalized HIV stigma predicts subsequent viremia in US HIV patients through depressive symptoms and antiretroviral therapy adherence. AIDS. 2020 Sep 1;34(11):1665–71.

9. Gamarel KE, Nelson KM, Stephenson R, Santiago Rivera OJ, Chiaramonte D, Miller RL, et al. Anticipated HIV stigma and delays in regular HIV testing behaviors among sexually-active young gay, bisexual, and other men who have sex with men and transgender women. AIDS Behav. 2018 Feb;22(2):522–30.

10. Ssenyonjo J, Shrestha R, Copenhaver M. Influence of stigma on pre-exposure prophylaxis (PrEP) care continuum among men and transwomen who have sex with men (MTWSM) in the United States. Int J HIV AIDS Prev Educ Behav Sci. 2019 Dec;5(2):134–40.

11. Lui CW, Dean J, Mutch A, Mao L, Debattista J, Lemoire J, et al. HIV testing in men who have sex with men: a follow-up review of the qualitative literature since 2010. AIDS Behav. 2018 Feb;22(2):593–605.

12. Babel RA, Wang P, Alessi EJ, Raymond HF, Wei C. Stigma, HIV risk, and access to HIV prevention and treatment services among men who have sex with men (MSM) in the United States: a scoping review. AIDS Behav. 2021 Nov;25(11):3574–604.

13. Nawfal ES, Gray A, Sheehan DM, Ibañez GE, Trepka MJ. A systematic review of the impact of HIV-related stigma and serostatus disclosure on retention in care and antiretroviral therapy adherence among women with HIV in the United States/Canada. AIDS Patient Care STDS. 2024 Jan;38(1):23–49.

14. Vermeer W, Gurkan C, Hjorth A, Benbow N, Mustanski BM, Kern D, et al. Agent-based model projections for reducing HIV infection among MSM: Prevention and care pathways to end the HIV epidemic in Chicago, Illinois. PLoS One. 2022;17(10):e0274288.

15. Probert WJM, Sauter R, Pickles M, Cori A, Bell-Mandla NF, Bwalya J, et al. Projected outcomes of universal testing and treatment in a generalised HIV epidemic in Zambia and South Africa (the HPTN 071 [PopART] trial): a modelling study. Lancet HIV. 2022 Nov;9(11):e771–80.

16. Abuelezam NN, McCormick AW, Surface ED, Fussell T, Freedberg KA, Lipsitch M, et al. Modelling the epidemiologic impact of achieving UNAIDS fast-track 90-90-90 and 95-95-95 targets in South Africa. Epidemiol Infect. 2019 Jan;147:e122.

17. Levy B, Correia HE, Chirove F, Ronoh M, Abebe A, Kgosimore M, et al. Modeling the effect of HIV/AIDS stigma on HIV infection dynamics in Kenya. Bull Math Biol. 2021 Apr 5;83(5):55.

18. Prudden HJ, Hamilton M, Foss AM, Adams ND, Stockton M, Black V, et al. Can mother-to-child transmission of HIV be eliminated without addressing the issue of stigma? Modeling the case for a setting in South Africa. PLOS ONE. 2017 Dec 8;12(12):e0189079.

19. Stover J, Glaubius R, Teng Y, Kelly S, Brown T, Hallett TB, et al. Modeling the epidemiological impact of the UNAIDS 2025 targets to end AIDS as a public health threat by 2030. PLOS Medicine. 2021 Oct 18;18(10):e1003831.

20. Campbell EM, Jia H, Shankar A, Hanson D, Luo W, Masciotra S, et al. Detailed Transmission Network Analysis of a Large Opiate-Driven Outbreak of HIV Infection in the United States. J Infect Dis. 2017 Nov 27;216(9):1053–62.

21. Bisanzio D, Reithinger R. Projected burden and duration of the 2022 Monkeypox outbreaks in non-endemic countries. The Lancet Microbe. 2022 Sep 1;3(9):e643.

22. Bisanzio D, Davis AE, Talbird SE, Van Effelterre T, Metz L, Gaudig M, et al. Targeted preventive vaccination campaigns to reduce Ebola outbreaks: An individual-based modeling study. Vaccine. 2023 Jan 16;41(3):684–93.

23. Bisanzio D, Reithinger R, Alqunaibet A, Almudarra S, Alsukait RF, Dong D, et al. Estimating the effect of non-pharmaceutical interventions to mitigate COVID-19 spread in Saudi Arabia. BMC Medicine. 2022 Feb 7;20(1):51.

24. Nyblade L, Mingkwan P, Stockton MA. Stigma reduction: an essential ingredient to ending AIDS by 2030. Lancet HIV. 2021 Feb 1;8(2):e106–13.

25. Golub SA, Gamarel KE. The impact of anticipated HIV stigma on delays in HIV testing behaviors: findings from a community-based sample of men who have sex with men and transgender women in New York City. AIDS Patient Care STDS. 2013 Nov;27(11):621–7.

26. Earnshaw VA, Smith LR, Chaudoir SR, Amico KR, Copenhaver MM. HIV stigma mechanisms and well-being among PLWH: a test of the HIV stigma framework. AIDS Behav. 2013 Jun;17(5):1785–95.

27. Pearson CA, Johnson MO, Neilands TB, Dilworth SE, Sauceda JA, Mugavero MJ, et al. Internalized HIV stigma predicts suboptimal retention in care among people living with HIV in the United States. AIDS Patient Care STDS. 2021 May;35(5):188–93.

28. Earnshaw VA, Chaudoir SR. From conceptualizing to measuring HIV stigma: a review of HIV stigma mechanism measures. AIDS Behav. 2009 Dec;13(6):1160–77.

29. Centers for Disease Control and Prevention. PrEP for HIV prevention in the U.S. [Internet]. 2023 [cited 2024 Apr 30]. Available from: https://web.archive.org/web/20240416105239/https://www.cdc.gov/nchhstp/newsroom/fact-sheets/hiv/PrEP-for-hiv-prevention-in-the-US-factsheet.html

30. HIV.gov. U.S. statistics [Internet]. 2024 [cited 2024 Apr 30]. Available from: https://www.hiv.gov/hiv-basics/overview/data-and-trends/statistics

31. AIDSVu. Understanding the current HIV epidemic in the United States [Internet]. [cited 2024 Apr 30]. Available from: https://map.aidsvu.org/profiles/nation/usa/overview#/

32. HIV.gov. U.S. statistics [Internet]. 2024 [cited 2024 Apr 30]. Available from: https://www.hiv.gov/hiv-basics/overview/data-and-trends/statistics

33. Lytvyn L, Siemieniuk RA, Dilmitis S, Ion A, Chang Y, Bala MM, et al. Values and preferences of women living with HIV who are pregnant, postpartum or considering pregnancy on choice of antiretroviral therapy during pregnancy. BMJ Open. 2017 Dec 1;7(9):e019023.

34. Kalichman SC, Simbayi LC, Cloete A, Mthembu PP, Mkhonta RN, Ginindza T. Measuring AIDS stigmas in people living with HIV/AIDS: the Internalized AIDS-Related Stigma Scale. AIDS Care. 2009 Jan;21(1):87–93.

35. Pantelic M, Boyes M, Cluver L, Thabeng M. “They say HIV is a punishment from God or from ancestors”: cross-cultural adaptation and psychometric assessment of an HIV stigma scale for South African adolescents living with HIV (ALHIV-SS). Child Indic Res. 2018;11(1):207–23.

36. Luseno WK, Field SH, Iritani BJ, Odongo FS, Kwaro D, Amek NO, et al. Pathways to Depression and Poor Quality of Life Among Adolescents in Western Kenya: Role of Anticipated HIV Stigma, HIV Risk Perception, and Sexual Behaviors. AIDS and Behavior [Internet]. 2020; Available from: 10.1007/s10461-020-02980-5

37. Kay ES, Rice WS, Crockett KB, Atkins GC, Batey DS, Turan B. Experienced HIV-Related Stigma in Health Care and Community Settings: Mediated Associations With Psychosocial and Health Outcomes. Journal of acquired immune deficiency syndromes (1999). 2018;77(3):257–63.

38. Stangl AL, Lloyd JK, Brady LM, Holland CE, Baral S. A systematic review of interventions to reduce HIV-related stigma and discrimination from 2002 to 2013: how far have we come? J Int AIDS Soc. 2013 Nov 13;16(3Suppl 2):18734.

39. Nyblade L, Mingkwan P, Stockton MA. Stigma reduction: an essential ingredient to ending AIDS by 2030. Lancet HIV. 2021 Feb 1;8(2):e106–13.

40. Mak WWS, Mo PKH, Ma GYK, Lam MYY. Meta-analysis and systematic review of studies on the effectiveness of HIV stigma reduction programs. Soc Sci Med. 2017 Sep;188:30–40.

41. Pantelic M, Steinert JI, Park J, Mellors S, Murau F. “Management of a spoiled identity”: systematic review of interventions to address self-stigma among people living with and affected by HIV. BMJ Glob Health. 2019;4(2):e001285.

42. Loutfy M, Tharao W, Logie C, Aden MA, Chambers LA, Wu W, et al. Systematic review of stigma reducing interventions for African/Black diasporic women. J Int AIDS Soc. 2015;18(1):19835.

43. Feyissa GT, Lockwood C, Woldie M, Munn Z. Reducing HIV-related stigma and discrimination in healthcare settings: A systematic review of quantitative evidence. PLoS One. 2019;14(1):e0211298.

44. Smith MK, Xu RH, Hunt SL, Wei C, Tucker JD, Tang W, et al. Combating HIV stigma in low- and middle-income healthcare settings: a scoping review. J Int AIDS Soc. 2020 Aug;23(8):e25553.

45. Aghaei A, Sakhaei A, Khalilimeybodi A, Qiao S, Li X. Impact of Mass Media on HIV/AIDS Stigma Reduction: A Systematic Review and Meta-analysis. AIDS Behav. 2023 Oct;27(10):3414–29.

46. Rueda S, Mitra S, Chen S, Gogolishvili D, Globerman J, Chambers L, et al. Examining the associations between HIV-related stigma and health outcomes in people living with HIV/AIDS: a series of meta-analyses. BMJ open. 2016;6(7):e011453.

47. Chan BT, Tsai AC. HIV stigma trends in the general population during antiretroviral treatment expansion: analysis of 31 countries in sub-Saharan Africa, 2003–2013. JAIDS Journal of Acquired Immune Deficiency Syndromes. 2016 Aug 15;72(5):558.

48. J Visser M. Change in HIV-related stigma in South Africa between 2004 and 2016: a cross-sectional community study. AIDS Care. 2018 Jun 3;30(6):734–8.

