## Supplemental file for "A novel modelling framework to simulate the effects of HIV stigma on HIV transmission dynamics"

### Model structure

The following section provides a detailed description of the HIV-IBM human social network components. Given the transmission dynamics of HIV, incubation time, and disease duration lag, the model runs on a weekly time step.

### Demographic data

The HIV spread simulation of the HIV-IBM was performed using a simulated population of 3 million people based on demographic statistics of the United States from the US Census Bureau (Table A1).[1]

*Table A1: Demographic parameters*

| Parameter | Value | Source |
|---|---|---|
| Average US household size | 2.6 | [1] |
| Age distribution | | [2] |
| < 18 | 22% | |
| 18-24 | 9.4% | |
| 25-44 | 26.3% | |
| 45-64 | 25.4% | |
| >65 | 16.8% | |
| Race | | [1] |
| Black or African American | 13.6% | |
| Not Black or African American | 86.4% | |
| Ethnicity | | [1] |
| Hispanic or Latino | 19.1% | |
| Not Hispanic or Latino | 81.9% | |
| Proportion of population who inject drugs | 1.5% | [3] |
| Proportion of PWID† who share needles | 32.1% | [4] |
| Sharing needle contacts | Mean 4.1 (standard deviation: 6.2 ) | [5] |

†People who inject drugs

### Human settlements

The model represented the spread of HIV in a city of 3 million people. We disaggregated the 3 million in-silico population in 226 cells as a 16x16 matrix, with each cell representing one square kilometre. Population density was 11,718.8 people per square kilometre, the mean population density of major cities in the US.[6] The population of each cell was estimated using a power distribution based on Zipf's law [7] fitted to a gridded US population obtained from the US Census Bureau.[1]

### Human sexual networks

#### *Modelling human sexual networks*

The HIV-IBM model structure was built to represent the high heterogeneity of sexual networks among people, as well as the "scale-free" and "small-world" characteristics common of many human social networks. In a "scale-free" network, most nodes (representing individuals) connect to a low number of other nodes, while a few have a high number of connections. The nodes with high connectivity are often called "super-spreaders." The link distribution among nodes in the scale-free network can be described by a power-law distribution:

$$p(x) = x^{-\alpha}$$

where $x$ is the number of links of a node, $p(x)$ is the cumulative distribution, and the exponent $\alpha$ is the scaling factor. For human social networks, the power-law distributions have exponent $\alpha$ values ranging from 2 to 3.[8] In a "small-world" network, two nodes in the network can reach each other through a short sequence of connected nodes (called a "short path").[9]

Sexual interactions among individuals are associated with many factors but have been shown to be well-described by networks with scale-free and small-world characteristics.[10,11]

#### *Creation of the HIV-IBM sexual network*

We built the sexual network of the synthetic population using information gathered from scientific literature. The sexual network was built to represent the following sexual orientations: women who have sex with men (WSM), men who have sex with women (MSW), women who have sex with women (WSW), men who have sex with men (MSM), women who have sex with both women and men (WSMSW), and man who have sex with both men and women (MSWSM). We assign each individual a sexual orientation based on reported proportions in the USA (Table A2). For each sexual orientation, we calculated the number of partners per person using a power law distribution with an *alpha* parameter set to have a mean number of partners equal estimates calculated using published data (Table A2). The model included the average weekly frequency of sexual activity for each sexually active individual based on annual data reported in Ueda et al [12].

*Table A2: Parameters used to build the synthetic population's sexual network*

| **Sexual orientation** | **Proportion of adult population (estimated)** | **Source** | **Mean number of partners per year** | **Source** | **Power law's alpha (estimated)** |
|---|---|---|---|---|---|
| WSM | 43.5% | [13] | 1.1 | [12,14] | 2.9 |
| MSW | 41.8% | [13] | 1.2 | [14] | 2.5 |
| WSW | 7.2% | [15–17] | 1.8 | [12,14] | 1.8 |
| MSM | 6.1% | [17–20] | 12.6 | [14,21] | 1.3 |
| WSMSW | 0.8% | [18–20] | 1.9 | [14] | 1.8 |
| MSWSM | 0.7% | [18–20] | 1.9 | [14] | 1.8 |

### Needles-sharing network among PWID

The model accounted for HIV transmission due to needle sharing among PWID. Thus, we created a network of needle sharing among PWID who share needles. The fraction of adult PWID among the synthetic population was set as 1.5% with 32.1% of them sharing needles (Table A1).[4] The distribution of needle-sharing contact among PWID sharing needles had a mean of 4.1 (standard deviation: 6.2).[5] We assumed that the sharing-needle followed a power-law distribution with $\alpha$=2.5 based on results shown in Campbell et al.[5]

### Human movement among city cells

The sexual and needle-sharing networks of the HIV-IBM also account for the movement of people by calculating a weight for each link based on the distance across cells and the estimated flux of people among cells. The weight was calculated using a gravity model accounting for distance between settlements and for population size.[22,23] A gravity model is a modified law of gravitation that, in its simpler formulation of a frictionless gravity model, considers the population size of two places and their distance apart to estimate the flow of people between them.[24] This method is not country-specific, but it has been used in West Africa to model Ebola spread.[23] The gravity model assumes that larger settlements attract more people and settlements closer together share more people than distant ones. The gravity model was created by merging population data and the distance matrix among the cells following the methods described in Balcan et al. and Kraemer et al.[22,23] The results of the gravity model were recorded in a flux matrix among cells, with columns and rows equal to the number of cells. Each cell of the flux matrix contained the estimated population flow between two cells.

### Modeling effect of stigma

The model objective is to investigate the impact of stigma on HIV transmission in a US-like synthetic population. The model seeks to represent the effects of different types of stigma experienced by people related to HIV infection. We simulate the effects of stigma using data from studies evaluating the level of stigma in a given population and its effects on HIV testing, HIV treatment start, and HIV treatment adherence. We focused on studies which reported stigma scales to quantify the level of stigma affecting the people included in the studies. Given that there is not a single standardized scale to measure different types of stigma, we used the scales adopted in the selected studies. The model simulated the effect of anticipated stigma on HIV testing and treatment adherence, internalised stigma on starting HIV treatment and treatment adherence, and enacted stigma on HIV treatment adherence (Table A3). We included the described stigma types based on the availability of studies providing enough data to assign stigma scores to each individual and calculate their effect on the probability of getting tested, starting treatment, and/or remaining adherent to treatment. We assigned a stigma score to all individuals per each scale used in the selected studies to represent stigma in the surveyed population. Thus, each individual had five independent stigma scores (Table A3). The weekly probability of getting tested, starting treatment after positive test, and remain adherent to treatment were calculated with the following formula

$$P = \left(\frac{1}{1+e^{-(\beta_0+\beta_1)}}\right)$$

Where *P* is the probability of the event, $\beta_1$ is the mean OR reported in the selected article, $\beta_0$ is a constant calculated to obtain the proportion of PLWH tested, under treatment, and adhering to treatment as reported in the Table XY for the baseline model. We performed three models to assess the effect of selected stigma typologies:

- *Baseline scenario*: stigma score per person was extracted from a normal distribution with mean and standard deviation equal to the those reported in the selected publications
- *Stigma halved scenario*: the scores assigned in the baseline models were reduced by 50% (stigma scores halved)
- *No stigma scenario*: the scores assigned in the baseline models were reduced by 100% (stigma scores set equal zero)

*Table A3: Parameters used to describe association among different types of stigmas and testing, starting treatment, and adherence in the HIV-IBM (all included models logistic regression)*

| **Stigma type** | **Affected behaviour** | **Mean stigma score** | **Standard deviation** | **Stigma effect (Odds ratio)** | **Source** |
|---|---|---|---|---|---|
| Anticipated stigma | Getting tested | 2.57 | 0.61 | 0.60 | [25] |
| Internalised stigma | Starting ART | 1.91 | 1.08 | 0.80 | [26] |
| Anticipated stigma | Adhering to ART | 1.93 | 0.70 | 0.38 | [27] |
| Internalised stigma | Not adhering to ART | 2.15 | 0.86 | 1.73 | [28] |
| Enacted stigma | Not adhering to ART | 1.31 | 0.46 | 1.38 | [28] |

### Modeling HIV epidemic

In the HIV-IBM, the infectious status of individuals followed the classical SEIR compartmental model structure: Susceptible (S) → Exposed (E) → Infectious (I) → Removed (R). Transition from one status to another was driven by use of HIV prevention technologies (pre-exposure prophylaxis (PrEP), condoms, and treatment as prevention), time from exposure, HIV care continuum stage, as well as sexual and needle sharing behaviours (Table A4). The model simulates the whole progression of the disease from exposure to symptoms.

*Table A4: HIV epidemiology parameters*

| Parameter | Point estimate | Range | Source |
|---|---|---|---|
| HIV transmission probability | | | |
| Per condomless sex act (base value) | 0.002 | | [29,30] |
| Per injection | 0.0067 | | [31] |
| Reduction in HIV transmission probability | | | |
| Due to condom use | 99.0% | | [32,33] |
| Due to PrEP† use | 99.0% | | [34] |
| If taking ART‡ and virally suppressed | 100.0% | | [35] |
| Time from exposure to virus detection | | | |
| Antibody/antigen test | | 18-90 days | [36] |
| Nucleic acid test | | 10-33 days | [36] |
| Time from exposure to onset of flu-like symptoms | | 14-28 days | [37] |
| Period of flu-like symptoms | | 7-14 days | [38] |
| Acute infection period | | 100-120 days | [39] |
| Peak of viremia | | 20-30 days | [39] |
| Increased sexual transmission | | 0.02 | [40] |
| Latency period from exposure to AIDS symptoms | | 10-15 years | [37,38] |
| Annual age-adjusted mortality rate | 12.1 per 1,000 people living with HIV | | [41] |

†PrEP: pre-exposure prophylaxis; ‡ART: anti-retroviral therapy

We started the simulation by seeding the in-silico population with 10,810 infected adults based on HIV prevalence in the USA (~3.6 infected per 1,000 people).[42] The seeding cases were proportionally distributed across sexual behaviour groups based on field surveys (Table A5).[42–44] The seeding cases represent individuals already living with HIV and who are at different HIV disease stages. The model accounts for the 10 fold increased transmission probability during the acute viremia period that occurs between 20-30 days after infection.[39,40] The model simulated sexual and injection-related transmission, which together account for more than 98% of all newly reported cases.[45] Weekly probabilities of HIV testing, initiating ART after positive test, and ART adherence were estimated at individual level.

*Table A5: HIV parameters used to create the starting status of the synthetic population*

| Parameter | Value | Source |
|---|---|---|
| PLWH† in the synthetic population | 10,810 | |
| By sexual orientation | | [42–44] |
| MSM | 6,831 | |

| | | |
|---|---|---|
| MSW | 972 | |
| MSWSM | 844 | |
| WSM | 2,053 | |
| WSW | 15 | |
| WSMSW | 1,798 | |
| Fraction of PLWH who are Black or African American | 38.3% | [42] |
| Fraction of PLWH who are Hispanic or Latino | 32.1% | [42] |
| Proportion of PLWH who know their status | 87.0% | [41] |
| Percentage of MSM and MSMSW annual screening rate | 35.3% | [46] |
| Percentage of MSW annual screening rate | 8.5% | [46] |
| Percentage of WSM/WSW/WSMSW annual screening rate | 21.0% | [47] |
| Proportion of people who test positive for HIV who start ART‡ within 1 month | 81.3% | [41] |
| Proportion diagnosed with HIV who are retained in care annually | 53.9% | [41] |
| Proportion diagnosed with HIV who are virally suppressed | 68.3% | [41] |
| Condom use: | | |
| MSM | 49.4% | [48] |
| MSW/WSM | 12.5% | [41] |
| PrEP coverage | 41.6% | [48] |

†PLWH people living with HIV; ‡ART: anti-retroviral therapy

### Model calibration

The baseline model was calibrated to obtain HIV incidence to have a confidence interval including the reported HIV incidence of the USA for the year 2022: 11.3 infected for every 100,000 people.[43] The calibration was obtained by changing the frequencies of weekly sexual activities of individuals by increasing or reducing the rate of sexual encounters.

### Sensitivity analyses

Sensitivity analyses were performed to assess the impact of changing stigma levels in the synthetic population on the HIV epidemic simulated in the HIV-IBM. The following input parameters were varied in the sensitivity analyses:

- Score scale of anticipated stigma affecting testing
- Score scale of slized stigma affecting starting ART treatment

- Score scale of anticipated stigma affecting starting treatment adherence
- Score scale of internalised stigma affecting starting treatment adherence
- Score scale of enacted stigma affecting starting treatment adherence

To test each parameter, we re-ran the HIV-IBM 10,000 times, randomly assigning a reduction of 0%, 50%, and 100% to each stigma score scale. The effect of each tested sigma scale on HIV transmission was then estimated using a generalized linear model (GLM) with a Gaussian link, where the GLM's dependent variable was the number of modeled HIV cases and the independent variables were the corresponding input stigma parameter sets [49].

### References


1. United States Census Bureau. QuickFacts: United States [Internet]. 2023 [cited 2024 Apr 24]. Available from: https://www.census.gov/quickfacts/fact/table/US/PST045222

2. Blakeslee L, Caplan Z, Meyer JA, Rabe MA, Roberts AW. Age and sex composition: 2020 [Internet]. United States Census Bureau; 2023 May [cited 2024 Apr 24]. (2020 Census Briefs). Report No.: C2020BR-06. Available from: https://www2.census.gov/library/publications/decennial/2020/census-briefs/c2020br-06.pdf

3. Bradley H, Hall EW, Asher A, Furukawa NW, Jones CM, Shealey J, et al. Estimated Number of People Who Inject Drugs in the United States. Clin Infect Dis Off Publ Infect Dis Soc Am. 2022 Jul 6;76(1):96–102.

4. Centers for Disease Control and Prevention. HIV infection risk, prevention, and testing behaviors among persons who inject drugs—national HIV behavioral surveillance: injection drug use, 23 U.S. cities, 2018 [Internet]. 2020 Feb [cited 2024 Apr 24]. Report No.: 24. Available from: https://www.cdc.gov/hiv/pdf/library/reports/surveillance/cdc-hiv-surveillance-special-report-number-24.pdf

5. Campbell EM, Jia H, Shankar A, Hanson D, Luo W, Masciotra S, et al. Detailed Transmission Network Analysis of a Large Opiate-Driven Outbreak of HIV Infection in the United States. J Infect Dis. 2017 Nov 27;216(9):1053–62.

6. United States Census Bureau. US 2020 Census [Internet]. Explore Census Data. [cited 2024 Apr 25]. Available from: https://data.census.gov/

7. Gabaix X. Zipf's law for cities: an explanation. Q J Econ. 1999;114(3):739–67.

8. Barrat A, Barthélemy M, Vespignani A. Dynamical processes on complex networks. 1st ed. Cambridge: Cambridge University Press; 2008.

9. Kleinberg J. Small-world phenomena and the dynamics of information. In: Advances in neural information processing systems. 2002. p. 431–8.

10. Hamilton DT, Handcock MS, Morris M. Degree distributions in sexual networks: a framework for evaluating evidence. Sex Transm Dis. 2008 Jan;35(1):30–40.

11. Liljeros F, Edling CR, Amaral LA, Stanley HE, Aberg Y. The web of human sexual contacts. Nature. 2001 Jun 21;411(6840):907–8.

12. Ueda P, Mercer CH, Ghaznavi C, Herbenick D. Trends in frequency of sexual activity and number of sexual partners among adults aged 18 to 44 years in the US, 2000-2018. JAMA Netw Open. 2020 Jun 12;3(6):e203833.

13. Flores AR, Conron KJ. Adult LGBT Population in the United States [Internet]. Los Angeles, CA: The Williams Institute, UCLA; 2023 Dec [cited 2024 Oct 1]. Available from: https://williamsinstitute.law.ucla.edu/publications/adult-lgbt-pop-us/

14. Mercer CH, Tanton C, Prah P, Erens B, Sonnenberg P, Clifton S, et al. Changes in sexual attitudes and lifestyles in Britain through the life course and over time: findings from the National Surveys of Sexual Attitudes and Lifestyles (Natsal). Lancet Lond Engl. 2013 Nov 30;382(9907):1781–94.

15. Tillewein H, Ma N. Implications for female same sex couples and HIV. Harv Public Health Rev [Internet]. 2022 Jun 4 [cited 2024 Sep 17];66. Available from: https://bcphr.org/66-article-ma/, https://bcphr.org/66-article-ma/

16. Przedworski JM, McAlpine DD, Karaca-Mandic P, VanKim NA. Health and health risks among sexual minority women: an examination of 3 subgroups. Am J Public Health. 2014 Jun;104(6):1045–7.

17. Jones JM. LGBTQ+ identification in U.S. now at 7.6% [Internet]. Gallup. 2024 [cited 2024 Sep 17]. Available from: https://news.gallup.com/poll/611864/lgbtq-identification.aspx.

18. Mauck DE, Gebrezgi MT, Sheehan DM, Fennie KP, Ibañez GE, Fenkl EA, et al. Population-based methods for estimating the number of men who have sex with men: a systematic review. Sex Health. 2019 Nov;16(6):527–38.

19. Lieb S, Fallon SJ, Friedman SR, Thompson DR, Gates GJ, Liberti TM, et al. Statewide estimation of racial/ethnic populations of men who have sex with men in the U.S. Public Health Rep. 2011 Jan 1;126(1):60–72.

20. Grey JA, Bernstein KT, Sullivan PS, Purcell DW, Chesson HW, Gift TL, et al. Estimating the population sizes of men who have sex with men in US states and counties using data from the American community survey. JMIR Public Health Surveill. 2016 Apr 21;2(1):e5365.

21. Grov C, Hirshfield S, Remien RH, Humberstone M, Chiasson MA. Exploring the venue's role in risky sexual behavior among gay and bisexual men: An event-level analysis from a national online survey in the U.S. Arch Sex Behav. 2013 Feb;42(2):291–302.

22. Balcan D, Colizza V, Gonçalves B, Hu H, Ramasco JJ, Vespignani A. Multiscale mobility networks and the spatial spreading of infectious diseases. Proc Natl Acad Sci. 2009 Dec 22;106(51):21484–9.

23. Kraemer MUG, Golding N, Bisanzio D, Bhatt S, Pigott DM, Ray SE, et al. Utilizing general human movement models to predict the spread of emerging infectious diseases in resource poor settings. Sci Rep. 2019 Mar 26;9:5151.

24. Anderson J. The gravity model. Annu Rev Econ. 2011;3(1):133–60.

25. Golub SA, Gamarel KE. The impact of anticipated HIV stigma on delays in HIV testing behaviors: findings from a community-based sample of men who have sex with men and transgender women in New York City. AIDS Patient Care STDs. 2013 Nov;27(11):621–7.

26. Pearson CA, Johnson MO, Neilands TB, Dilworth SE, Sauceda JA, Mugavero MJ, et al. Internalized HIV stigma predicts suboptimal retention in care among people living with HIV in the United States. AIDS Patient Care STDs. 2021 May;35(5):188–93.

27. Shrestha R, Altice FL, Copenhaver MM. HIV-related stigma, motivation to adhere to antiretroviral therapy, and medication adherence among HIV-positive methadone-maintained patients. J Acquir Immune Defic Syndr 1999. 2019 Feb 1;80(2):166–73.

28. Earnshaw VA, Smith LR, Chaudoir SR, Amico KR, Copenhaver MM. HIV stigma mechanisms and well-being among PLWH: a test of the HIV stigma framework. AIDS Behav. 2013 Jun;17(5):1785–95.

29. Vitinghoff E, Douglas J, Judon F, McKiman D, MacQueen K, Buchinder SP. Per-Contact Risk of Human Immunodificiency Virus Tramnsmision between Male Sexual Partners. Am J Epidemiol. 1999 Aug 1;150(3):306–11.

30. Patel P, Borkowf CB, Brooks JT, Lasry A, Lansky A, Mermin J. Estimating per-act HIV transmission risk: a systematic review. AIDS. 2014 Jun 19;28(10):1509–19.

31. Kaplan EH, Heimer R. A model-based estimate of HIV infectivity via needle sharing. JAIDS J Acquir Immune Defic Syndr. 1992 Nov;5(11):1116.

32. Weller S, Davis K. Condom effectiveness in reducing heterosexual HIV transmission. Cochrane Database Syst Rev. 2002;(1):CD003255.

33. Smith DK, Herbst JH, Zhang X, Rose CE. Condom effectiveness for HIV prevention by consistency of use among men who have sex with men in the United States. J Acquir Immune Defic Syndr 1999. 2015 Mar 1;68(3):337–44.

34. Centers for Disease Control and Prevention. PrEP effectiveness [Internet]. 2022 [cited 2024 Apr 25]. Available from: https://www.cdc.gov/hiv/basics/prep/prep-effectiveness.html

35. Rodger AJ, Cambiano V, Bruun T, Vernazza P, Collins S, van Lunzen J, et al. Sexual activity without condoms and risk of HIV transmission in serodifferent couples when the HIV-positive partner is using suppressive antiretroviral therapy. JAMA. 2016 Jul 12;316(2):171–81.

36. Centers for Disease Control and Prevention. Understanding the HIV window period [Internet]. 2022 [cited 2024 Apr 25]. Available from: https://www.cdc.gov/hiv/basics/hiv-testing/hiv-window-period.html

37. Centers for Disease Control and Prevention. About HIV [Internet]. 2022 [cited 2024 Apr 25]. Available from: https://www.cdc.gov/hiv/basics/whatishiv.html

38. National Health Services. Overview: HIV and AIDS [Internet]. 2021 [cited 2024 Apr 25]. Available from: https://www.nhs.uk/conditions/hiv-and-aids/

39. McMichael AJ, Borrow P, Tomaras GD, Goonetilleke N, Haynes BF. The immune response during acute HIV-1 infection: clues for vaccine development. Nat Rev Immunol. 2010 Jan;10(1):11–23.

40. Pilcher CD, Tien HC, Eron JJ, Vernazza PL, Leu SY, Stewart PW, et al. Brief but efficient: acute HIV infection and the sexual transmission of HIV. J Infect Dis. 2004 May 15;189(10):1785–92.

41. Centers for Disease Control and Prevention. Monitoring selected national HIV prevention and care objectives by using HIV surveillance data—United States and 6 dependent areas, 2019 [Internet]. Atlanta, Georgia; 2021 May [cited 2024 Apr 25]. (HIV Surveillance Supplemental Report). Report No.: 26(No.2). Available from: https://www.cdc.gov/hiv/library/reports/hiv-surveillance/vol-26-no-2/index.html

42. Centers for Disease Control and Prevention. PrEP for HIV prevention in the U.S. [Internet]. 2023 [cited 2024 Apr 30]. Available from: https://web.archive.org/web/20240416105239/https://www.cdc.gov/nchhstp/newsroom/fact-sheets/hiv/PrEP-for-hiv-prevention-in-the-US-factsheet.html

43. HIV.gov. U.S. statistics [Internet]. 2024 [cited 2024 Apr 30]. Available from: https://www.hiv.gov/hiv-basics/overview/data-and-trends/statistics

44. AIDSVu. Understanding the current HIV epidemic in the United States [Internet]. [cited 2024 Apr 30]. Available from: https://map.aidsvu.org/profiles/nation/usa/overview#/

45. Centers for Disease Control and Prevention. HIV Surveillance Report: Diagnoses, Deaths, and Prevalence of HIV in the United States and 6 Territories and Freely Associated States, 2022. 2024 May 21; Available from: https://stacks.cdc.gov/view/cdc/156509

46. Spensley CB, Plegue M, Seda R, Harper DM. Annual HIV screening rates for HIV-negative men who have sex with men in primary care. PLOS ONE. 2022 Jul 14;17(7):e0266747.

47. Chandra A, Billioux VG, Copen CE, Balaji A, DiNenno E. HIV testing in the U.S. household population aged 15-44: data from the National Survey of Family Growth, 2006-2010. Natl Health Stat Rep. 2012 Oct 4;(58):1–26.

48. Centers for Disease Control and Prevention. HIV infection risk, prevention, and testing behaviors among men who have sex with men—national HIV behavioral surveillance, 13 U.S. cities, 2021 [Internet]. 2023 Jan [cited 2024 Apr 25]. Report No.: 31. Available from: https://www.cdc.gov/hiv/library/reports/hiv-surveillance-special-reports/no-31/index.html

49. Bisanzio D, Davis AE, Talbird SE, Van Effelterre T, Metz L, Gaudig M, et al. Targeted preventive vaccination campaigns to reduce Ebola outbreaks: An individual-based modeling study. Vaccine. 2023 Jan 16;41(3):684–93.
